# Meta-analysis study of the therapeutic impact of Mesenchymal stem cells derived exosomes for chronic kidney diseases

**DOI:** 10.1101/2024.09.04.24313060

**Authors:** k Himanshu, k Gunjan, Ramendera Pati Pandey, Riya Mukherjee, Chung-Ming Chang

## Abstract

Mesenchymal stem cell-derived exosomes (EXOs) represent a promising avenue for treating chronic kidney diseases (CKD), though their precise impact remains somewhat elusive. To address this gap, we conducted a systematic analysis, scouring databases and clinical trial repositories for relevant studies from 2019 to 2023. Seventeen papers were meticulously selected for their focus on mesenchymal stem cell-derived exosomes (MSC-EXOs) and their potential in CKD treatment. Our comprehensive meta-analysis, incorporating 15 preclinical and 6 clinical studies, underscores the efficacy of MSC-EXOs in improving renal function while attenuating tubular injury, inflammation, apoptosis, collagen deposition, and renal fibrosis. Notably, post-treatment with MSC-EXOs exhibited significant associations with various CKD markers, with pooled proportions indicating a considerable impact on blood urea nitrogen (BUN) and serum creatinine (SCR) levels. Subgroup analyses based on animal models further elucidated heterogeneity within the studies. In conclusion, MSC-EXOs demonstrate promise in enhancing renal function and reducing CKD risk, as evidenced by both preclinical and clinical data. Their efficacy in lowering SCR and BUN levels while enhancing filtration rate suggests MSC-EXOs as a viable and secure alternative to cell-based therapies, thereby providing valuable insights for personalized CKD treatments despite inherent limitations.

**Graphical abstract:** 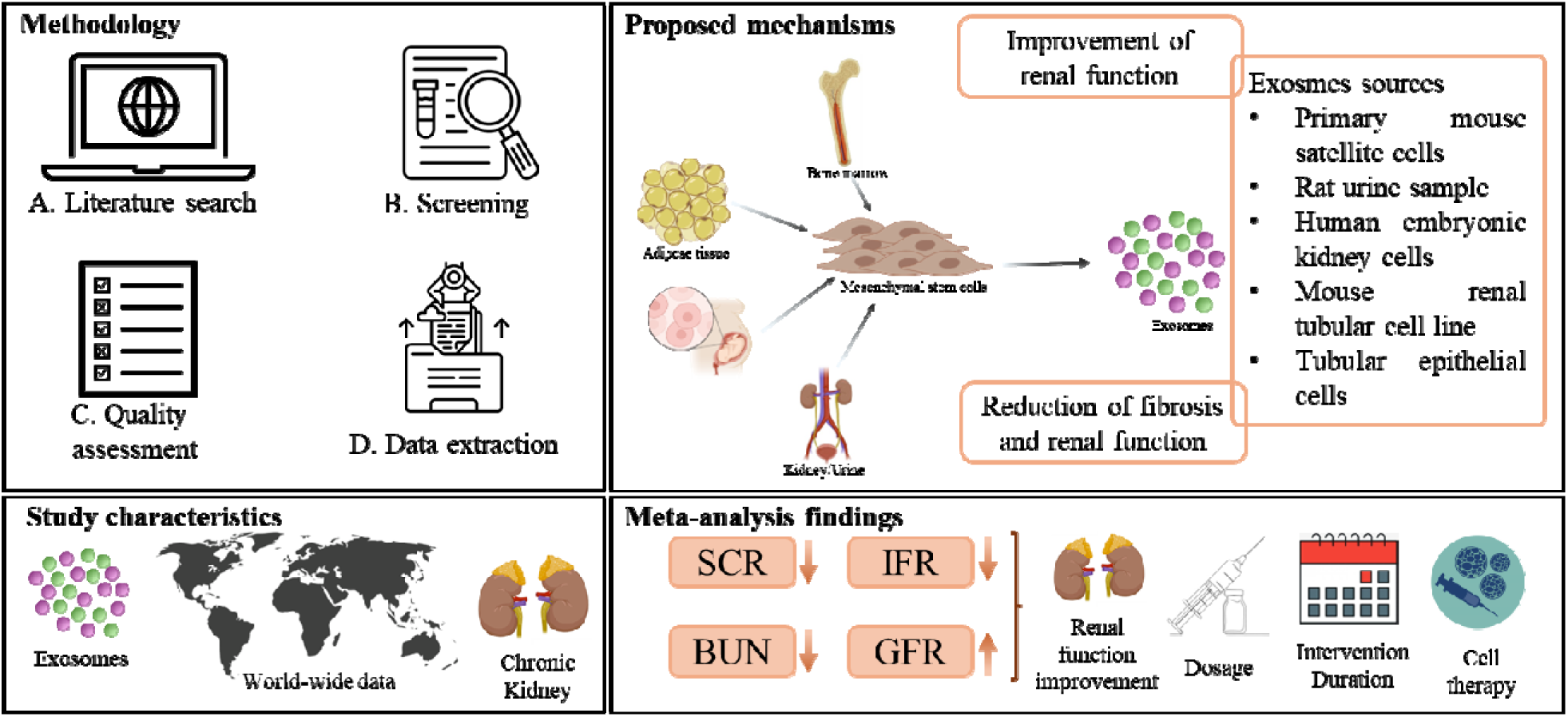

Graphical highlight:

1. MSC-EXOs show promise in improving renal function and reducing CKD risk, as evidenced by comprehensive meta-analysis findings.
2. Pooled proportions indicate significant associations between MSC-EXO treatment and reduced BUN and SCR levels and increased GFR rate, highlighting their potential therapeutic efficacy.
3. Exosomes show a significant effect on renal function, as indicated by the study findings.

## Introduction

Chronic kidney disease (CKD) emerges as a significant global health concern that demands attention. It is elevated incidence, substantial healthcare burden, insidious onset, poor prognosis, and other associated challenges pose serious threats to the well-being of individuals worldwide[1, 2]. CKD emerges because of diminished oxygen delivery to the kidneys. Subsequent amplification of kidney hypoxia leads to compromised regenerative capacity, cell damage, oxidative stress, eliciting inflammatory responses and renal fibrosis within the kidney compartments[3]. To counteract these processes, numerous pharmaceutical therapies have been developed. There are five phases of CKD, and each one is linked to increased risks of cardiovascular morbidity, early mortality, and/or a poor standard of life[4]. Renal damage, indicative of CKD, encompasses pathological abnormalities, abnormal urine sediment, or an elevated urinary albumin excretion rate, detectable through imaging or renal biopsy[5]. Currently, none of these therapies have been clinically validated to effectively alter the outcome of CKD. [6–8]. By the year 2040, it is projected to become the fifth most prevalent cause of death globally[9]. Unfortunately, conventional pharmacological methods often overlook the intricate interconnections and complexities of overlapping disease-related mechanisms[10]. An alternate approach including targeted delivery and control of disease pathways through the transfer facilitated by extracellular vesicles (EVs) could address the bottleneck between regenerative medicine and current pharmaceutical treatments.[11].

According to recent data, CKD is a global health issue, affecting approximately 9% to 13% of the worldwide population (approximately 700 million to one billion people), with a significant portion of patients falling into stage 3 of the disease.[4, 5, 13]. It rates up to 15% in the USA, that is about 37 million people, causing a high economic burden[13]. Tragically, millions of individuals die each year due to the lack of affordable treatment options. The 2023 ISN-GKHA report highlights the widespread impact of CKD, with around 850 million people affected globally, regardless of age[14]. Disadvantaged populations are particularly vulnerable to the disease. The high costs of treatment and the significant health consequences associated with kidney disease contribute to its devastating effects. In Taiwan, CKD has become a major concern, ranking as the 9th leading cause of death over the past decade[15]. Research estimates that the national prevalence of CKD in Taiwan is approximately 11.9%, affecting more than 2.5 million people[16, 17]. Furthermore, Taiwan has a more than 1.5-fold greater prevalence of end-stage renal disease (ESRD) than the United States, with approximately 3400 individuals per 1 million in the general population experiencing kidney failure[18]. These statistics highlight the urgent need for improved access to affordable treatment options and increased awareness about CKD worldwide.

In recent years, cell-based therapies have garnered attention across various medical research fields. Mesenchymal stromal cells (MSCs) are extracted from diverse adult tissues, including adipose tissue, umbilical cord blood, bone marrow, and macrophage[19] MSCs are multipotent cells that can differentiate into tissues derived from mesoderm and have the ability to self-renew. [20]. Furthermore, MSC possesses the ability to evade alloantigen recognition owing to their low immunogenicity and lack of expression of co-stimulatory molecules. The inherent immunomodulatory properties of MSCs, coupled with their minimal potential side effects, present a therapeutic alternative in this regard[21]. Recent studies suggest that administration of MSC-derived EVs ameliorates CKD in preclinical models. [22–25].

In the realm of renal diseases, EXO derived from MSC have gained significant attention due to their pathophysiological, diagnostic, and therapeutic roles [26]. These nanosized vesicles, ranging from 30 to 100 nm in diameter, originate from multivesicular bodies [27]. Under both physiological and pathological conditions, various cell release EXO into the blood or other bodily fluids, reflecting cellular responses to internal and external stimuli [28]. EXO carry a diverse cargo, including proteins, long noncoding RNAs, microRNAs (miRNAs), mRNAs, and lipids of particular note, miRNAs, which are noncoding, single-stranded small RNAs, which play a vital part in regulating gene expression by binding to the 3’ untranslated regions (UTRs) of target mRNAs, which leads to disintegration or translational suppression. [29, 30]. Preclinical studies have showcased the positive impacts of MSC-derived EVs ameliorates CKD derived from cells, including secreted growth factors, microvesicles, and EXO, in models of chronic kidney injury[31]. These findings suggest a regenerative influence of cell-based therapies on renal function. Additionally, MSCs are actively employed in several clinical trials involving kidney transplant recipients, with the goal of enhancing immunosuppression and promoting improved regeneration [32]. Additionally, these EXO are free from the adverse aspects related to tumorigenic and immunogenic associated with cellular therapies, making them a safer and more viable avenue for future regenerative medicine [33].

In this meta-analysis and systematic review aimed to assess the impact of MSC derived EXO on outcome parameters related to chronic kidney disease function and morphology, scrutinizing both cell- and model-related aspects. The existing studies can enhance the design of future clinical investigations. Additionally, the insights gained can be utilized to refine current experimental animal models and interventions, thereby improving the quality of preclinical research in the future.

## Result

### Study selection and characteristics

A total of 758 papers published from 2019 to 2023 were systematically categorized using esteemed databases, including PubMed (n = 186), Web of Science (n = 95), Google Scholar (n = 402), and Cochrane (n = 56), along with 19 additional records sourced from other sources. Out of these categorized articles 427 duplicate articles were removed, within this selected, 331 articles 224 articles were excluded due to reasons such as their classification as review articles (89), conference papers (17), case reports (76), book chapters (26), and abstracts (16). Our meticulous screening process meticulously refined the dataset to 107 studies suitable for our systematic review. This subset underwent further scrutiny, resulting in the exclusion of 94 articles. These exclusions were attributed to diverse reasons, including publications in languages other than english (3), inappropriate study designs (38), and absence of extracellular vesicle (EV) administration in treatment (23), unsuitable outcomes (7), and unrepresentative results (19). This exhaustive evaluation ultimately led to the inclusion of 17 pre-clinical studies [34–50] that met our stringent inclusion criteria. The study selection process adhered to the PRISMA[51] flow diagram, as shown in figure 1.

**Figure 1:**
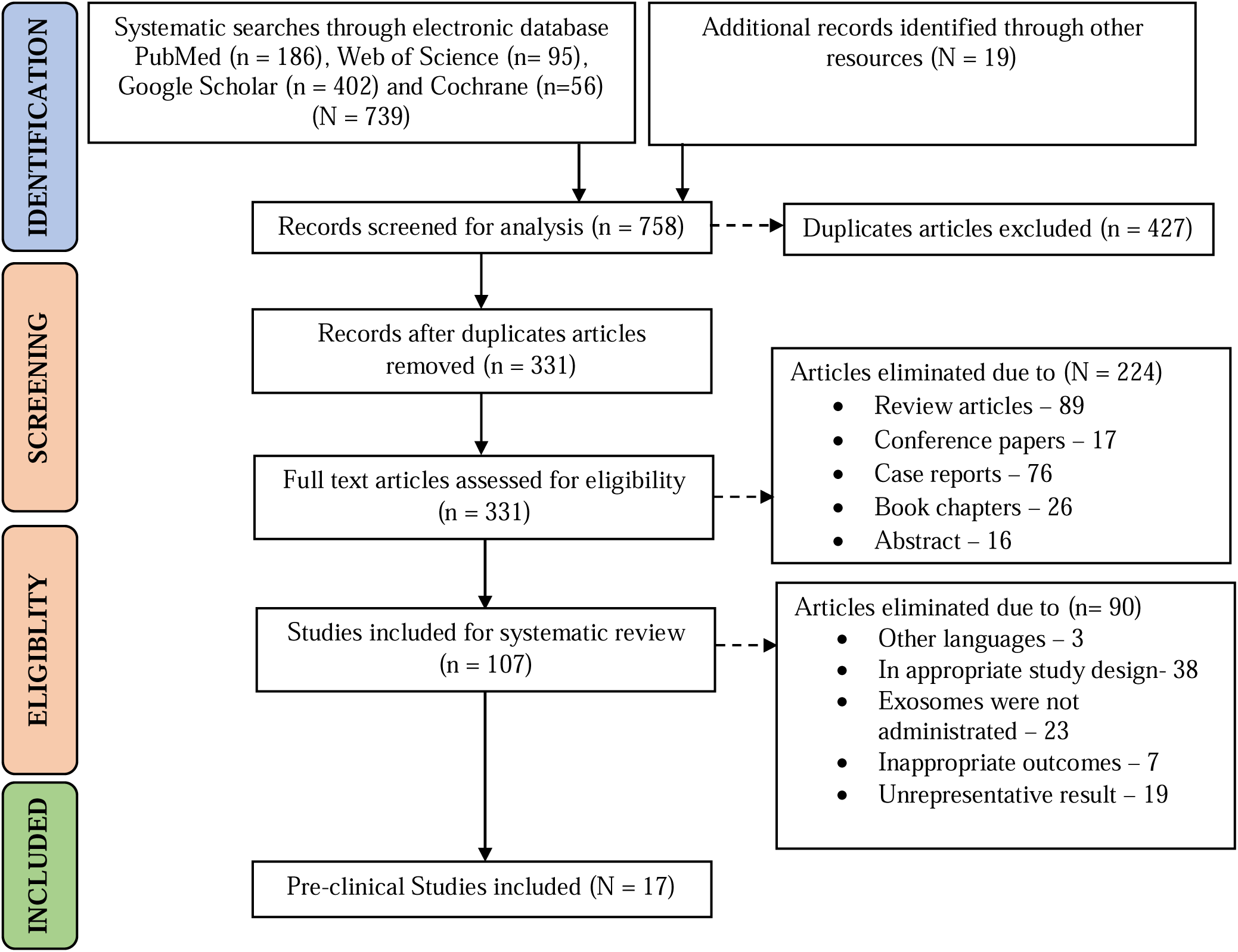
Study selection process according to PRISMA.

### Quality assessment

The risk of bias assessment for the included studies reveals variations in specific domains. Xi Liu et al [44], Yingjie Liu et al. [36] and H. Wang et al.[52] is deemed to have a high risk of incomplete outcome data (attrition bias), indicating potential concerns regarding the completeness of outcome data reported in this study. Conversely, M. Liang et al. [53], and Yan Wang et al. [42] and H. Wang et al. [52] are characterized by a low risk of baseline characteristics and other bias, suggesting that the methods used to assess and measure outcomes in these studies are robust and unlikely to introduce bias, as shown in figure 2.

**Figure 2:**
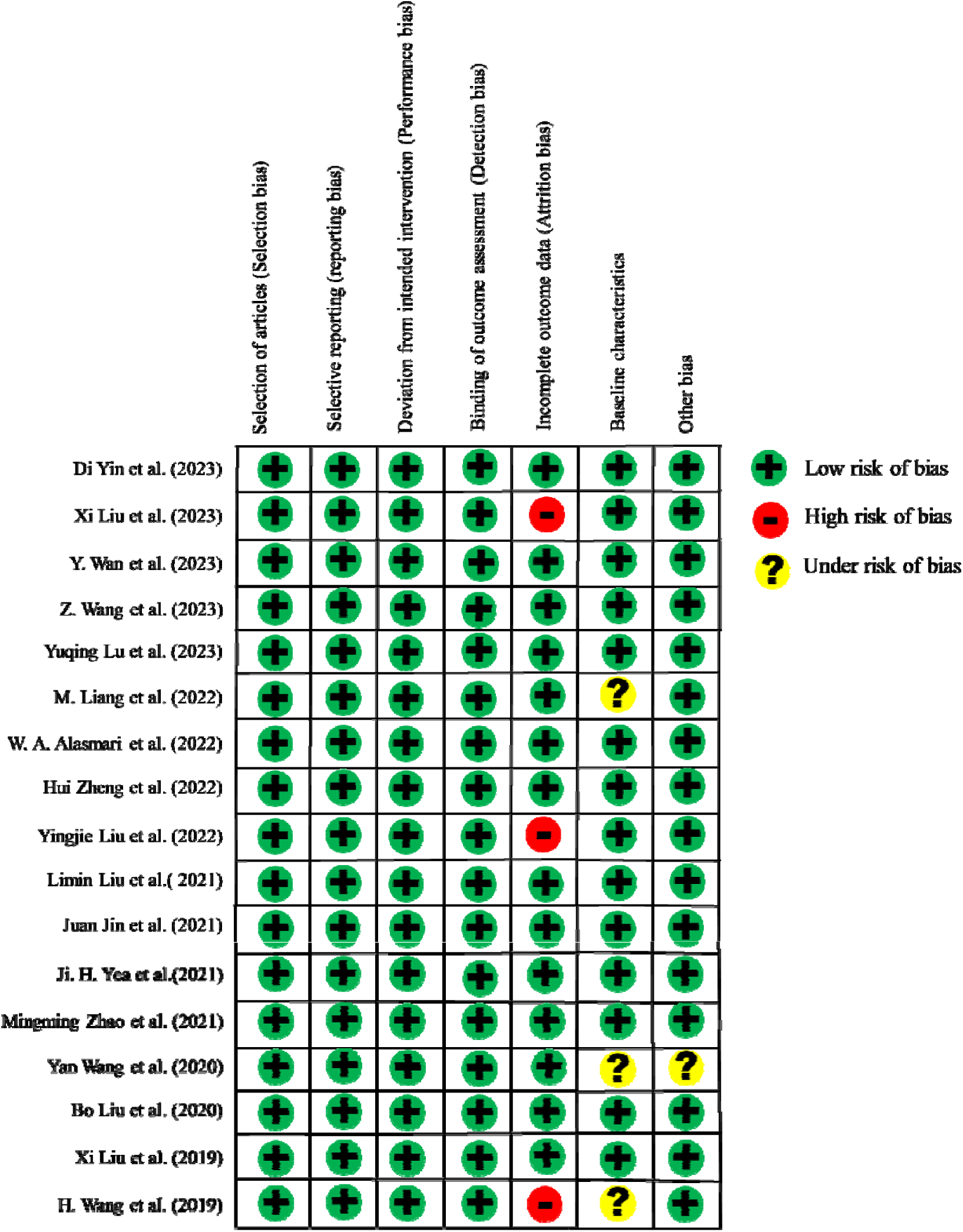
Risk of BIAS

## General characteristics

### Clinical

CKD is a multifaceted and advancing health issue, often requiring a range of treatments and approaches. One area of promising exploration involves EXO, which has emerged for their potential therapeutic impact on CKD. To determine the efficacy and safety of employing EXO-based therapies for CKD, numerous clinical trials have been carried out. These trials encompass a spectrum of EXO sourced from different origins, such as MSCs, bone marrow, adipose tissue, umbilical cord, and various other cell types. The diversity in exosome sources is aimed at understanding their distinct compositions and potential roles in alleviating CKD-related complications.

To the best of our current knowledge, there are no registered clinical trials for the treatment of CKD using exosomes on clinicaltrials.gov. However, we have compiled a list of ongoing clinical trials that explore the therapeutic potential of MSCs for diverse medical conditions. These trials exhibit a varied landscape in terms of clinical status, phases, locations, study types, sources of EXO, and study durations. Among the six trials with known clinical status, one has been completed (NCT02166489), four are actively recruiting (NCT05512988, NCT03939741), and NCT05362786 is active but not recruiting, while the status of the remaining two is unknown (NCT03321942, NCT05042206). These trials encompass both allogeneic and autologous approaches, with three employing allogeneic MSCs, three using autologous MSCs, and one unspecified. In terms of phases, three trials are in Phase I, two are in Phase I/II, and one does not specify its phase. Geographically, the trials are spread across different countries, including one in the United States, China, Iran, Korea, and Bangladesh. The MSC used in these trials primarily originates from bone marrow (n =3), followed by adipose tissue (n = 2) and umbilical cord (n = 1), as summarized in table 1.

**Table 1:** Clinical trials of MSCs in treating chronic kidney diseases. *Abbreviations:* N/A: not applicable; I.V.: intravenous

| S. No | Id no. | Location | Source | Autologous/allogeneic | Study type | Phase | Number of patients | Administration frequency | Start year | End year | Status | Outcome measures |
| --- | --- | --- | --- | --- | --- | --- | --- | --- | --- | --- | --- | --- |
| 1. | NCT05362786 | United States | Bone marrow-derived mesenchymal stem cell | Allogenic | Non-randomized, open label | Phase I | 14 | I.V. injection, delivered MSC in one of two fixed dosing regimens at two time points | 2022 | N/A | Active, not recruiting | - Primary: adverse events and/or serious adverse events and change in Gfr value |
| 2. | NCT05512988 | China | Umbilical cord derived mesenchymal stem cells (UC-MSC) | Autologous | Randomized, double-blind, controlled clinical study | Phase I phase II | 44 | IV injection, each trial group received two injections throughout the entire course, with a cumulative dose of $2 \times 10^6$ cells/kg/person. | 2021 | N/A | Recruiting | N/A |
| 3. | NCT03321942 | China | Adipose-derived mesenchymal stem/stromal cells (MSC) | N/A | Randomized, open label | N/A | 100 | Intravenous injection for 3 months | 2017 | N/A | Unknown status | - Primary: mass formation & creatinin<br>- Secondary: Gfr |
| 4. | NCT02166489 | Iran | Bone marrow-derived mesenchymal stem cell | Autologous | Open label | Phase I | 6 | I.V. injection of high doses ($2 \times 10^6$) of autologous MSC per Kg of their body weight, sourced from bone marrow biopsies. | 2014 | 2016 | Completed | - Primary: mass formation<br>- Secondary: Gfr |
| 5. | NCT05042206 | Korea | Allogeneic bone marrow derived mesenchymal stem | Allogeneic | Open label, single center | Phase I | 10 | I.V. injection of 10ml dose 3 times in 2 weeks. | 2021 | N/A | Unknown status | - Primary: adverse event<br>- Secondary: changes in EGfr, bun and creatinine |
| 6. | NCT03939741 | Bangladesh | Adipose derived stem cell (ADSC) | Autologous | Randomized, double-blind, controlled, open label | Phase I/ phase II | 31 | I.V. injection of 5ml stromal vascular fraction (SVF) containing ADSC. | 2019 | N/A | Recruiting | - Primary: change of Gfr & adverse events,<br>- Secondary: Iga, igg, igm, c3, c4, crp, il-6, peripheral hemolymocyte subsets. |

### Pre-clinical

This meta-analysis and systematic review meticulously explore exosome-based therapeutic interventions for chronic kidney disease (CKD) through a comprehensive analysis of diverse sources. The study encompasses data from various animal models, including C57BL/6 (n=9), SD rats (n=4), BALB/c (n=2), Albino rats (n=1), and Bred Fisher rats (n=1), originating from countries such as the USA (n=2), China (n=13), Saudi Arabia (n=1), and South Korea (n=1). The investigation includes a gender-specific breakdown, focusing on human (n=11) and animal (n=17) subjects, and incorporates a variety of cell types, such as human umbilical cord MSC (hucMSCs) (n=3), human bone marrow-derived MSCs (BM-MSC) (n=4), adipose tissue-derived MSC (n=1), among others. Diverse exosome isolation methods are examined, comprising ultracentrifugation (n=8), centrifugation + filtration (n=7), and exosome isolation kits (n=2). Characterization techniques involve nanoparticle tracking analysis (NTA) (n=2), transmission electron microscopy (TEM) (n=5), both TEM and NTA (n=9), and N/A (n=1). The study identifies a consistent EXO size range of 30 to 150 nm, with common markers CD9, CD63, and TSG101. Administration routes vary, including intravenous (n=14), intramuscular (n=1), intraperitoneal (n=1), and intracellular (n=1), with concentrations ranging from 20 mg to 250 μg, providing a thorough overview of EXO -based therapies in managing CKD. At the culmination of this analysis, these findings underscore the diverse landscape of EXO -based therapies in addressing CKD, offering valuable insights into the potential efficacy and methodologies employed across various studies, aiming to elucidate optimal strategies for leveraging EXO therapy in CKD management, as summarized in table 2.

**Table 2:**
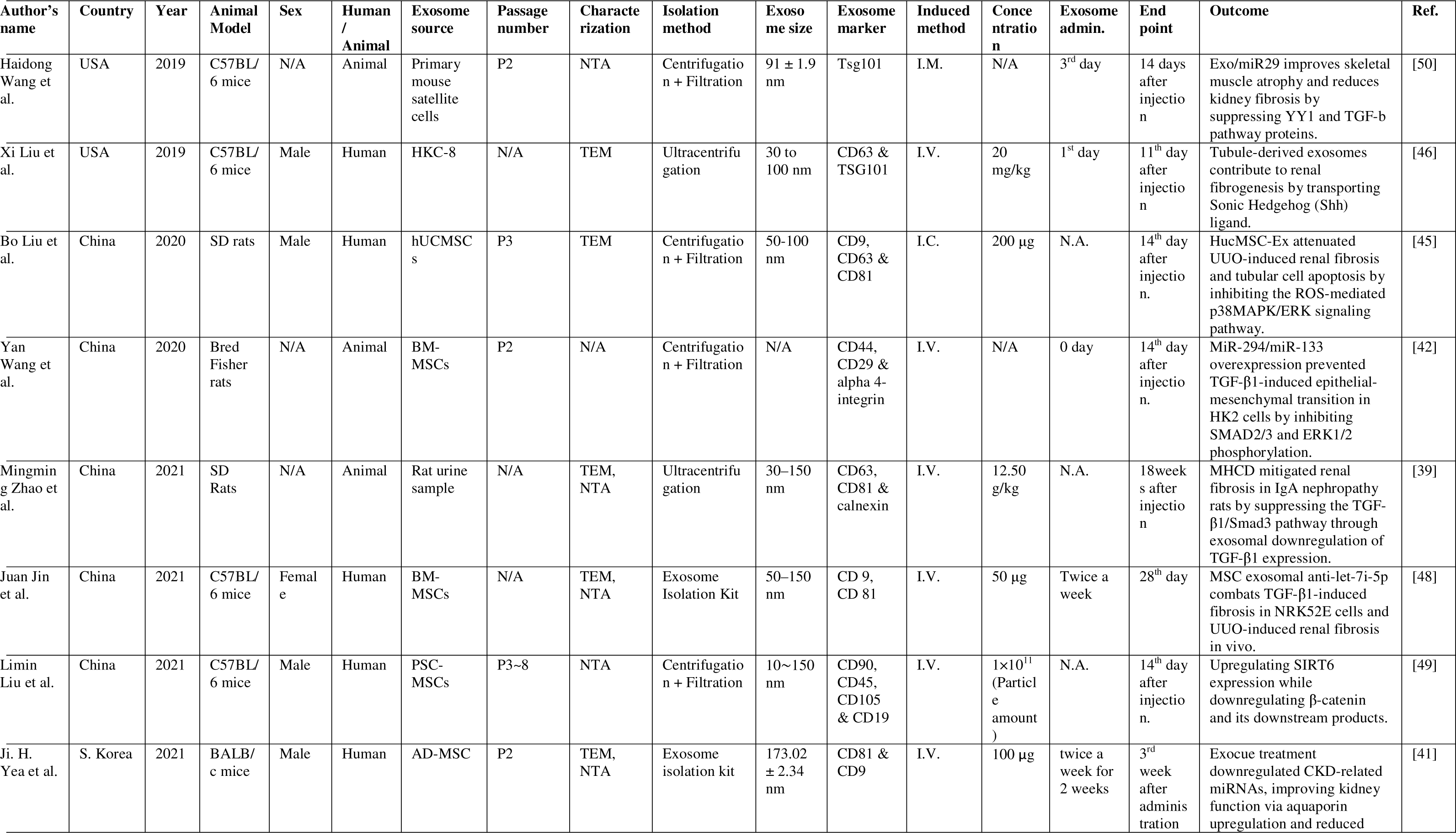
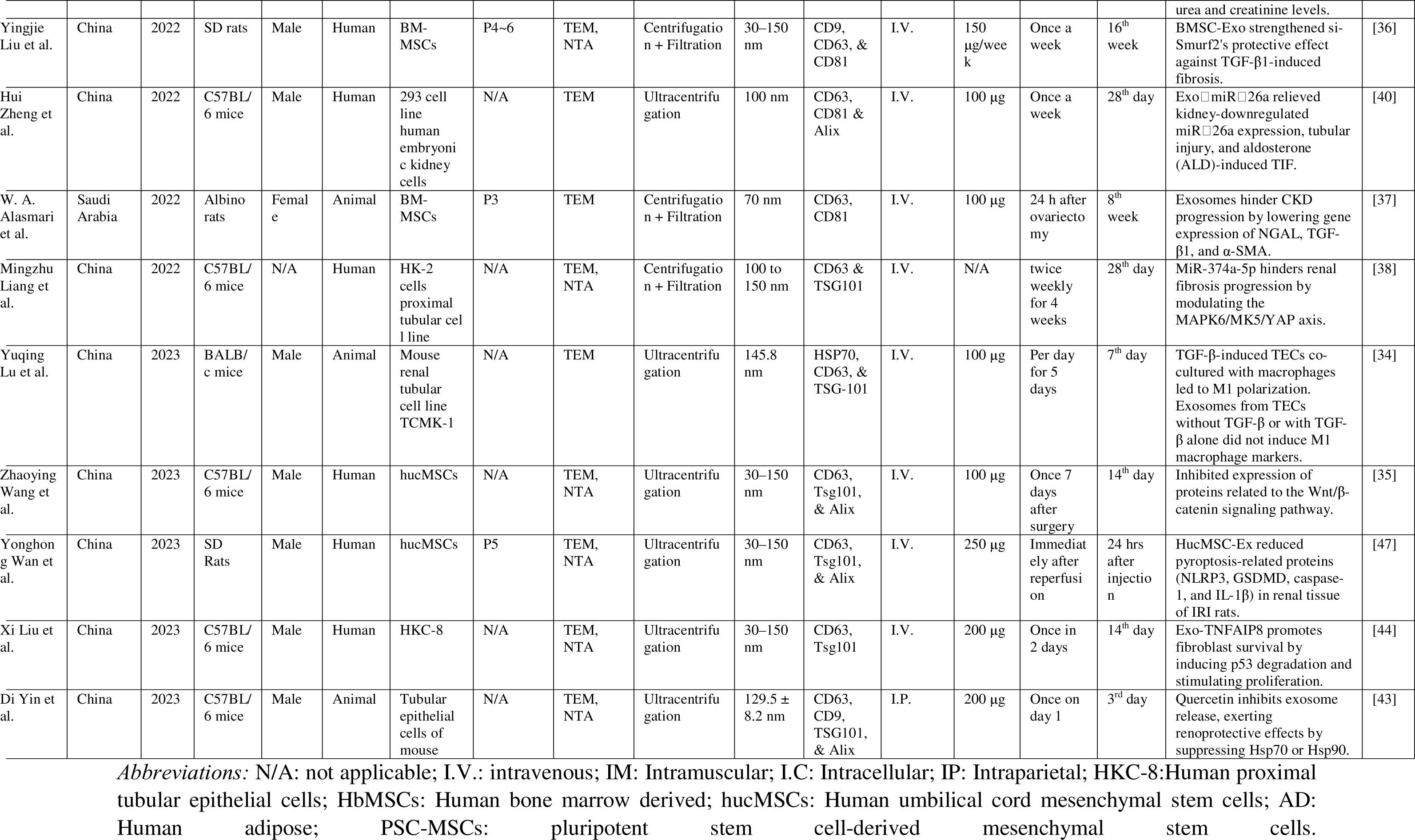
Characteristics of included preclinical studies for meta-analysis.

## Meta-analysis

### A. Primary outcome

#### i. Blood urea nitrogen (BUN)

BUN is a waste product that is produced when protein is broken down in the body[54]. The analysis of BUN in CKD reveals a notable degree of heterogeneity, as indicated by an I^2^ value of 92%, *τ*^2^ of 0.0573%, and a significant p-value of less than 0.01. This substantial heterogeneity underscores the diversity in BUN outcomes across the studies included in the review. The calculated proportion and its 95% confidence interval (CI) further emphasize this variability, with a value of 0.55 [0.41; 0.68], as depicted in figure 3A.

**Figure 3:**
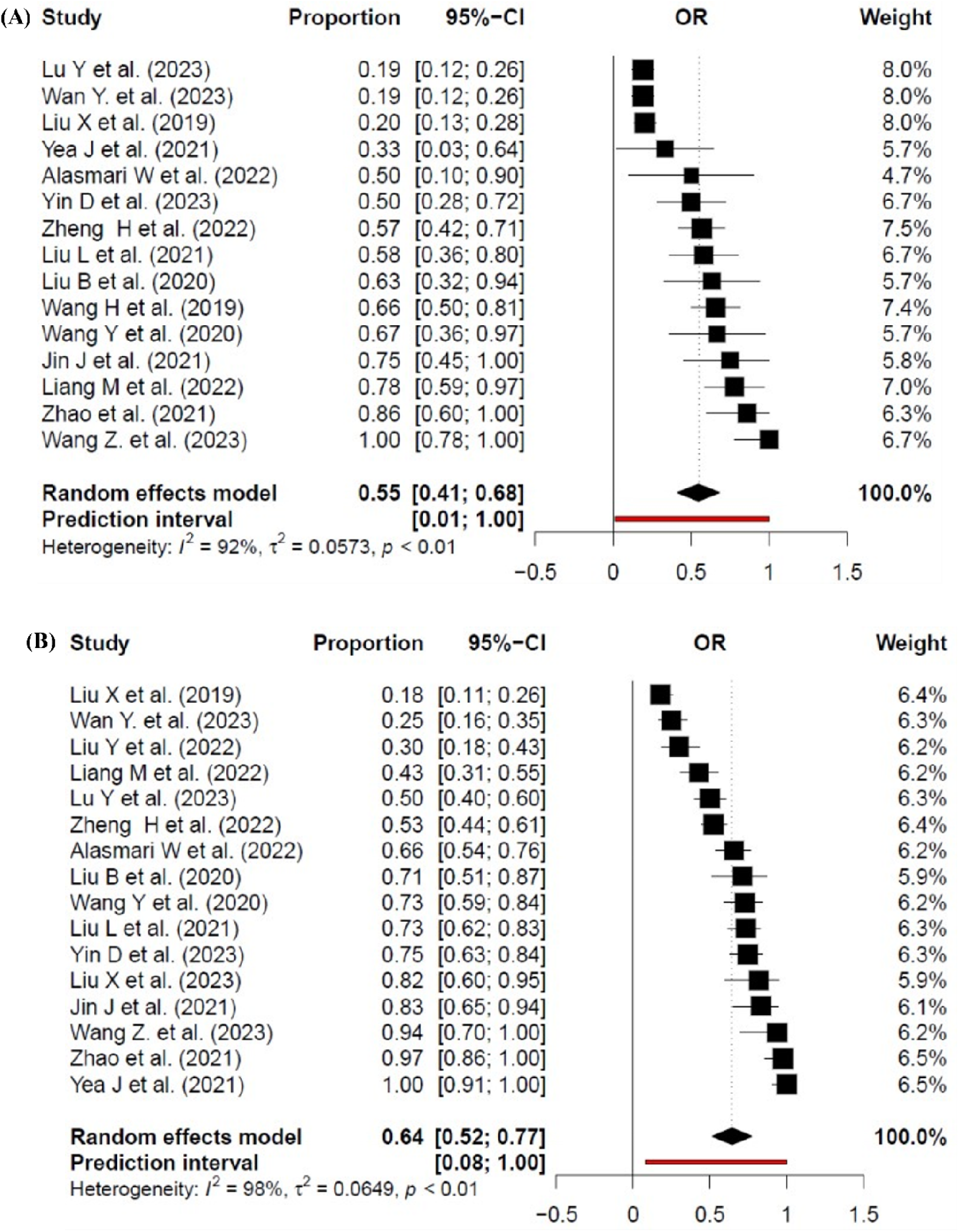
BUN and SCR level expressed with their 95% confidence intervals (A) efficacy in BUN reduction of MSC derived EXOs in the CKD model (B) efficacy in SCR reduction of MSC derived EXOs in the CKD model.

#### ii. Serum creatinine (SCR)

SCR is a waste product that is produced when creatinine, a chemical that is found in muscle tissue, is broken down[54]. In the context of CKD, the analysis of SCR levels reveals a high degree of heterogeneity, as evidenced by an I^2^ value of 98%, *τ*^2^ of 0.0649%, and a significant p-value of less than 0.01. This substantial heterogeneity underscores the varied outcomes observed across studies included in the analysis. The calculated proportion and its 95% CI further emphasize this diversity, with the value of 0.64 [0.52; 0.77], as depicted in figure 3B.

### B. Secondary outcome

#### i. Animal model subgroup analysis based on BUN level

In the evaluation of BUN in animal models of CKD, notable heterogeneity is observed across various species. Among C57BL/6 mice, there is a significant degree of heterogeneity with an I^2^ value of 93%, *τ*^2^ of 0.0504%, and a p-value less than 0.01. The proportion estimate, with its 95% CI), is calculated at 0.62 [0.45; 0.79], with this animal model contributing 56.0% of the overall weight. Similarly, SD rats exhibit high heterogeneity (I^2^: 93%, *τ*^2^: 0.1122%, p <0.01), with a proportion estimate of 0.54 [0.14; 0.94] and contributing 20.0% to the overall analysis. Bred Fisher rats, on the other hand, demonstrate a proportion estimate of 0.67 [0.36; 0.77], representing 5.6% of the total weight. BALB/c mice display substantial heterogeneity (I^2^: 86%, *τ*^2^: 0.0820%, p <0.01), with a proportion estimate of 0.58 [0.15; 1.00] and contributing 13.8% to the overall analysis. Albino rats exhibit a proportion estimate of 0.59 [0.47; 0.72], accounting for 4.6% of the total weight. In aggregate, there remains significant heterogeneity across all models (I^2^: 94%, *τ*^2^: 0.0495%, p <0.01), with an overall proportion estimate of 0.59 [0.47; 0.72], as depicted in figure 4A.

**Figure 4:**
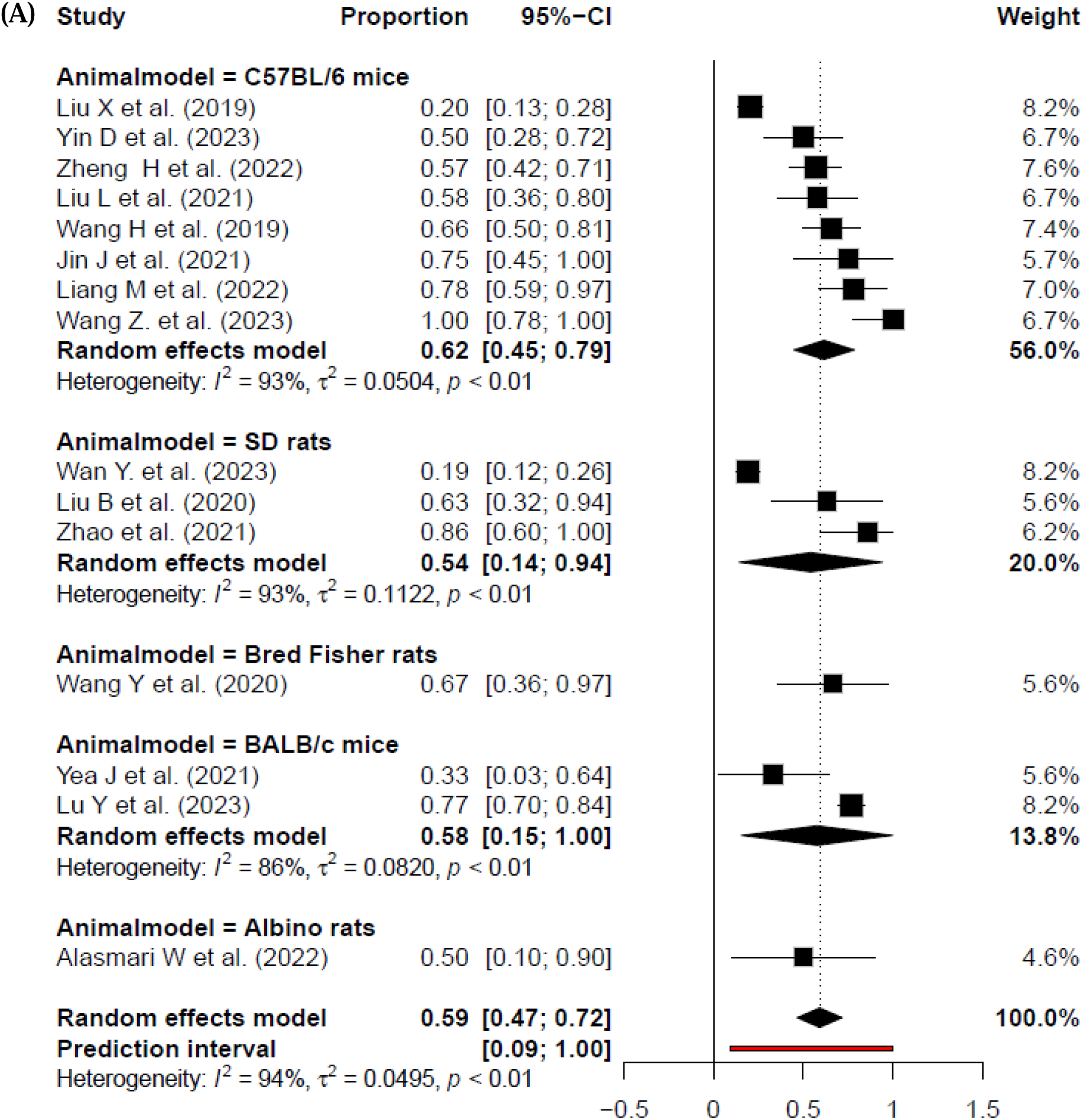
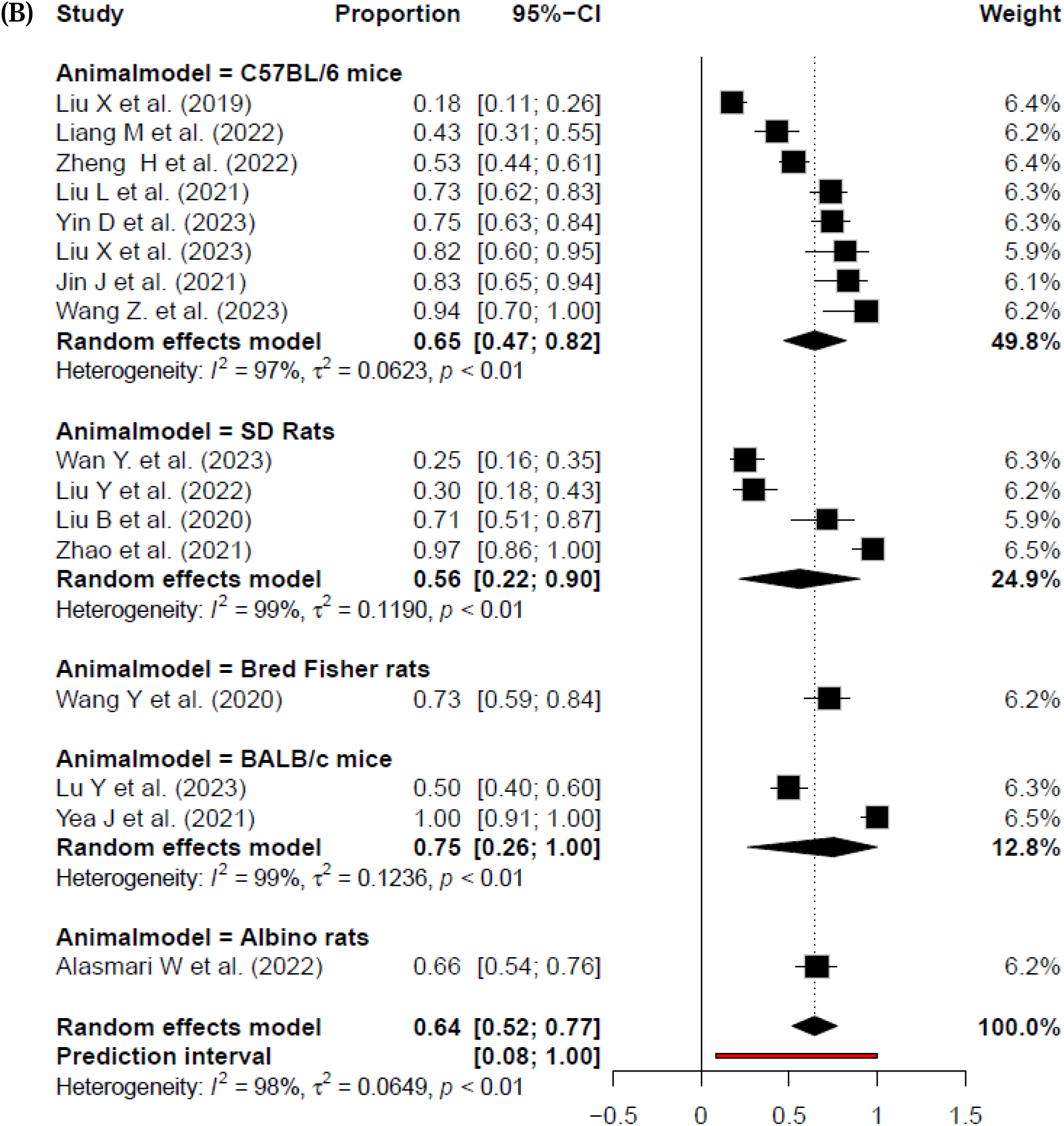
Subgroup analysis based on animal model with their 95% confidence intervals (A) efficacy in BUN reduction of MSC derived EXOs (B) efficacy in SCR reduction of MSC derived EXOs.

#### ii. Animal model subgroup analysis based on SCR level

In the investigation of SCR levels across various animal models of CKD, significant heterogeneity is observed. Among C57BL/6 mice, a high degree of heterogeneity is evident with an I^2^ value of 97%, tau square of 0.0623%, and a p-value less than 0.01. The proportion estimate, accompanied by its 95% CI, is determined to be 0.65 [0.47; 0.82], with this animal model contributing 49.8% to the overall weight. Similarly, SD rats display pronounced heterogeneity (I^2^: 99%, *τ*^2^: 0.1190%, p <0.01), with a proportion estimate of 0.56 [0.22; 0.90] and representing 24.9% of the total weight. Bred Fisher rats, in contrast, exhibit a proportion estimate of 0.73 [0.59; 0.84], contributing 6.2% to the overall analysis. BALB/c mice demonstrate substantial heterogeneity (I^2^: 99%, *τ*^2^: 0.1236%, p <0.01), with a proportion estimate of 0.75 [0.26; 1.00] and contributing 12.8% to the overall weight. Albino rats display a proportion estimate of 0.66 [0.52; 0.76], accounting for 6.2% of the total weight. Collectively, significant heterogeneity persists across all models (I^2^: 98%, *τ*^2^: 0.0649%, p <0.01), with an overall proportion estimate of 0.64 [0.52; 0.77], as depicted in figure 4B.

## Discussion

In this comprehensive systematic review and meta-analysis, encompassing 17 preclinical and 6 clinical studies, we provide a thorough examination of the impact of MSC-EXOs on diverse models of CKD. Preclinical and clinical studies have demonstrated that treating patients with CKD using MSC-EXOs leads to reductions in SCR and BUN levels, improved glomerular filtration rate, and protection of renal functions, along with the suppression of inflammatory responses. Though our knowledge of the distinct parts of vesicular structures, their specific functions, and their roles as therapeutic vectors, biomarkers, and contributors to autoimmune disorders is still insufficient. The exploration of EXOs in CKD pathogenesis is still in its early stages, with limited research compared to other diseases. Further research is essential for the clinical application of MSC-EXOs. Although the safety of MSC-EXOs has been initially confirmed through animal experiments, extensive studies are required to ensure their safety for clinical use. Various MSC sources, such as bone marrow, adipose tissue and umbilical cord, contribute to EXOs derivation, necessitating additional experiments to assess the safety of different MSC-EXOs.

The delivery of EXOs has various benefits as a cell-free therapeutic approach, such as enhanced stability, reduced immunogenicity, permeability, and cytotoxicity[55]. As a result, employing EXOs could offer a practical and secure substitute for cell-based treatments. Many studies attribute these beneficial effects primarily to the RNA cargo carried by EVs, and it’s noteworthy that these effects can be abrogated by RNase treatment. Numerous miRNA candidates, such as miR29, miR-294/miR-133, miR□26a, and miR-374a-5p, have been implicated in the pathophysiological processes of CKD. Research has revealed specific mechanisms through which various miRNAs contribute to mitigating kidney fibrosis and related complications in CKD. For instance, miR29 has been shown to ameliorate skeletal muscle atrophy and diminish kidney fibrosis by suppressing YY1 and proteins involved in the TGF-β pathway [50]. Additionally, by blocking the phosphorylation of SMAD2/3 and ERK1/2, miR-294/miR-133 has shown promise in preventing TGF-β1-induced epithelial-mesenchymal transition in HK2 cells [42]. Moreover, downregulation of miR□26a expression has been associated with reduced tubular injury and tubulointerstitial fibrosis induced by aldosterone[40]. Furthermore, miR-374a-5p has been shown to impede the progression of renal fibrosis by modulating the MAPK6/MK5/YAP axis, ultimately leading to reductions in urea and creatinine levels [38].

Beyond the influence of microRNAs, various therapeutic interventions involving MSCs and their derivatives have demonstrated promising outcomes in combating renal fibrosis and CKD. For instance, EXO derived from tubules play a role in promoting renal fibrogenesis by transporting the Sonic Hedgehog ligand [46]. HucMSC-ExO have demonstrated effectiveness in reducing renal fibrosis induced by UUO and preventing tubular cell apoptosis by inhibiting the ROS-mediated p38MAPK/ERK signaling pathway [45]. Additionally, it has been demonstrated that modified EXO employing heparin-chitosan MHCD reduces renal fibrosis in rats suffering from IgA nephropathy by inhibiting the TGF-β1/Smad3 pathway by lowering TGF-β1 expression in the EXO cargo [39]. Moreover, anti-let-7i-5p treatment with BM-MSCs has been effective in countering TGF-β1-induced fibrosis in NRK52E cells and inhibiting UUO-induced renal fibrosis *in vivo* [48]. PSC-MSCs-derived EXO have shown the ability to upregulate SIRT6 expression while downregulating β-catenin and its downstream products [49]. Additionally, BM-MSC-derived EXO have enhanced the protective effects against TGF-β1-induced fibrosis when combined with si-Smurf2 [36]. BM-MSCs EXOs have further demonstrated their impact on hindering CKD progression by lowering the gene expression of NGAL, TGF-β1, and α-SMA [37]. In co-culture experiments, TGF-β-induced tubular epithelial cells with macrophages led to M1 polarization, whereas BM-MSC EXO from TECs without TGF-β or with TGF-β alone did not induce M1 macrophage markers [34]. HucMSC-derived EXOs have shown a reduction in pyroptosis-related proteins (IL-1β, NLRP3, caspase-1, and GSDMD) in renal tissue of rats with ischemia-reperfusion injury (IRI) [35]. Furthermore, EXOs from HKC-8 cells, when treated with TNFAIP8, promoted fibroblast survival by inducing p53 degradation and stimulating proliferation [44, 47]. Additionally, in mouse tubular epithelial cells, quercetin has been identified to inhibit EXOs release, exerting reno-protective effects by suppressing Hsp70 or Hsp90 [43]. These diverse strategies involving MSCs-EXO and their derivatives showcase their potential in addressing various aspects of renal fibrosis and CKD progression.

Understanding the specific study design and factors influencing the efficacy of EXOs based treatments can enhance the planning of future experimental studies and aid in designing studies for specific patient populations. Accordingly, for studies with available creatinine (SCR) and urea data (BUN), we conducted uni-variable stratified meta-analyses to explore potential predictors for EXO-based therapy effectiveness across diverse CKD settings. To elucidate the impact of EXO treatment on various animal models, we conducted subgroup analysis graphs. Our findings indicate that the functional efficacy of EXO therapy varies depending on the model employed. Notably, a significant proportion of the animal records analyzed in our study pertained to CKD models.

As a cell-free therapeutic approach, EXO present numerous advantages, characterized by high stability and permeability, as well as low immunogenicity and cytotoxicity [56]. This suggests that the administration of EXOs could serve as a viable and safe alternative to cell-based therapies. Despite this potential, it’s important to acknowledge substantial heterogeneity in the literature, and even after conducting subgroup analysis, this heterogeneity persisted.

In summary, MSC-EXOs demonstrate promise in improving renal function and mitigating the risk of CKD. Our findings, supported by both preclinical and clinical evidence, confirm the efficacy of MSC-EXOs in lowering SCR and BUN levels, enhancing glomerular filtration rate, and alleviating renal inflammation within a rodent model of CKD. This study suggests that EXOs could serve as a secure alternative to cell-based therapies. The prospects involve addressing current challenges, conducting clinical trials, standardizing protocols, and advancing our understanding of EXO-based therapies to pave the way for their effective clinical implementation in the management of CKD. This work paves the way for future studies to contribute to the development of advanced therapies for CKD and underscores the potential for growth and innovation in the biopharmaceutical sector

## Materials and methods

### a. Literature search

For our present study, we conducted a comprehensive search across multiple databases, including Google scholar, PubMed/MEDLINE, Scopus, EMBASE, and the Web of Science, CINAHL, the Cochrane Register for Controlled Trials (CENTRAL), LILACS, SciELO, etc. with a focus on studies related to EXO-mediated treatment of CKD. The latest search update was performed on January 31, 2024. The search strategy involved employing various combinations of keywords, such as (“extracellular vesicle” OR “EV” OR “exosome”) AND (“stromal cell” OR “stem cell” OR “SC” OR “mesenchymal stem cell” OR “MSC”) and (“kidney disease” OR “kidney dysfunction” OR “chronic kidney disease” OR “renal Fibrosis” OR “Chronic Renal Failure” OR “chronic renal insufficiency” OR “CKD” OR “renal dysfunction”). We assessed study quality using defined criteria from the Collaborative Approach to Meta-analysis and Review of Animal Data in Experimental Studies (CAMRADES) risk of bias checklist. Moreover, for clinical studies, we conducted a thorough data search utilizing a comprehensive database encompassing privately and publicly funded clinical trials conducted globally, available at https://clinicaltrials.gov/. Additionally, manual searches of bibliographies and reference lists were performed to identify any additional relevant studies. No ethical approval was required as the meta-analysis relied solely on published articles.

### b. Inclusion criteria

Following the elimination of duplicates, the articles were screened using the predetermined inclusion and exclusion criteria. Inclusion criteria encompassed: (1) original articles; (2) animal experiments utilizing rats, mice etc.; (3) focus on chronic kidney fibrosis or renal fibrosis; (4) intervention involving the injection of EVs derived from any MSC source; (5) EXOs obtained from either stem cells or pluripotent cells, with a specific emphasis on CKD, and mandated the inclusion of at least one group subjected to EXO treatment.; (6) preconditioned, and modified MSC-EXOs (such as those transfected with genes or featuring overexpression of proteins or microRNAs) were encompassed in the study. (7) administration of MSC-EXOs involved either allogeneic, xenogeneic or autologous approaches. and (8) assessment of therapeutic efficacy as the outcome.

### c. Exclusion criteria

Exclusion criteria involved: (1) studies lacking a control group or involving inappropriate comparisons; (2) those not focusing on CKD models; (3) research combining therapeutic approaches with other agents of uncertain effects; and (4) article types such as comments, letters, reviews, editorials, and case reports, and (5) animal model used for other kidney related disorders, such as acute kidney disease, stroke and kidney stone formation.

### d. Data extraction

Data extraction from all eligible studies encompassed gathering the following information for the clinical trials table: authors’ names, study location, status of the trials, study year, type of kidney disease, study type, phase, patients’ number, autologous/allogeneic, administration, frequency, outcome measures and intervention findings. Similarly, the preclinical trials table was compiled using the following details: author’s name, country, publication year, animal model and their sex, human or animal derived, EXO source, isolation methods, modified strategy, and outcomes. Additionally, another data was compiled in a table by using information such as: author’s name, characterization techniques, EXO size, induction methods, concentration, time of EXO administration and end point of the study. Spreadsheets were created to facilitate the extraction and synthesis of the data by using Excel® (Microsoft® Office Excel 2021) and subjected to pre-testing before complete extraction. Citations from the compiled papers were managed using Mendeley software (version 2.105.0, Elsevier, London, UK).

### e. Quality assessment

Assessing publication bias is crucial to ensuring the integrity and credibility of the meta-analysis focused on the effects of EXOs on various aspects of CKD. To gauge the potential impact of publication bias in our findings, we applied several established techniques widely recommended in the field. A key method involved visually inspecting a bias risk graph for asymmetry, which may indicate the presence of publication bias. By employing these comprehensive approaches, our goal was to systematically address any potential bias and guarantee that our meta-analysis offers an unbiased synthesis of the current evidence regarding the positive effects of EXOs in the context of CKD.

### f. Statistical analysis

The study compiled data on CKD, EXO source, and group sizes from papers or correspondence with authors. Employing random-effects models in meta-analysis, individual effects were considered, avoiding fixed effects to effectively address unobserved heterogeneity. Results were presented using effect size and 95% CI. The metafor package in R (https://www.R-project.org/) facilitated all analyses. Notably, the meta-analysis specifically targeted SCR and BUN levels within the study.

## Data Availability

All data produced in the present work are contained in the manuscript

## Ethics approval and consent to participate

’Not applicable’

## Consent for publication

All authors have given their consent to publish

## Availability of data and materials

The authors confirm that the data supporting the findings of this study are available within the article.

## Authors’ contributions

All authors have equally contributed to the conceptualization, methodology, and writing and editing of the manuscript.

## Acknowledgements

This research was funded by VtR Inc-CGU (SCRPD1L0221); DOXABIO-CGU (SCRPD1K0131), and CGU grant (UZRPD1L0011, UZRPD1M0081). We have not used AI in the preparation of the manuscript.

